# Location of ryanodine receptor type 2 mutation predicts age of onset of sudden death in catecholaminergic polymorphic ventricular tachycardia – A systematic review and meta-analysis of case-based literature

**DOI:** 10.1101/2024.03.15.24304349

**Authors:** Halil Beqaj, Leah Sittenfeld, Alexander Chang, Marco Miotto, Haikel Dridi, Gloria Willson, Carolyn Martinez Jorge, Jaan Altosaar Li, Steven Reiken, Yang Liu, Zonglin Dai, Andrew R. Marks

**Author notes:** These authors contributed equally (Halil Beqaj, Leah Sittenfeld).

## Abstract

**Background:** Catecholaminergic polymorphic ventricular tachycardia (CPVT) is a rare inherited arrhythmia caused by mutations in the ryanodine receptor type 2 (RyR2). Diagnosis of CPVT often occurs after a major cardiac event, thus posing a severe threat to the patient’s health.

**Methods:** Publication databases, including PubMed, Scopus, and Embase, were searched for articles on patients with RyR2-CPVT mutations and their associated clinical presentation. Articles were reviewed by two independent reviewers and mutations were analyzed for demographic information, mutation distribution, and therapeutics. The human RyR2 cryo-EM structure was used to model CPVT mutations and predict the diagnosis and outcomes of CPVT patients.

**Findings:** We present a database of 1008 CPVT patients from 227 papers. Data analyses revealed that patients most often experienced exercise-induced syncope in their early teenage years but the diagnosis of CPVT took a decade. Mutations located near key regulatory sites in the channel were associated with earlier onset of CPVT symptoms including sudden cardiac death.

**Interpretation:** The present study provides a road map for predicting clinical outcomes based on the location of RyR2 mutations in CPVT patients. The study was partially limited by the inconsistency in the depth of information provided in each article, but nevertheless is an important contribution to the understanding of the clinical and molecular basis of CPVT and suggests the need for early diagnosis and creative approaches to disease management.

**Funding:** The work was supported by grant NIH R01HL145473, P01 HL164319 R25HL156002, T32 HL120826.

## Introduction

Catecholaminergic polymorphic ventricular tachycardia (CPVT) is a malignant arrhythmia associated with stress- and exercise-induced syncope and sudden cardiac death (SCD)^1^ in the absence of cardiac structural abnormalities^2^. The mortality rates are 30-50% by the age of 40^3^. Thus, early recognition, risk stratification, and intervention as indicated are crucial to achieving good outcomes for CPVT patients.

This rare inherited arrhythmia syndrome is caused by mutations in the cardiac ryanodine receptor (RyR2) or the sarcoplasmic reticulum protein calsequestrin 2 (CASQ2) gene^4–6^. RyR2 mutations are responsible for the autosomal dominant form of CPVT and account for approximately 60% of clinically well-defined CPVT^7^. RyR2 is a homotetrameric intracellular calcium release channel in cardiac muscle that mediates the release of calcium from the sarcoplasmic reticulum (SR) and plays a key role in excitation-contraction coupling (ECC)^8^. During increased stimulation of beta-adrenergic receptors of the heart (due to exercise or stress), RyR2 is phosphorylated by protein kinase A (PKA) at Ser2808^9^. This phosphorylation depletes the stabilizing subunit calstabin2^10^, increasing both the sensitivity of RyR2 to calcium-dependent activation and the open probability of the channel, and thus placing the entire channel structure into a “primed state”^11^. The primed state leads to more readily activated channels at low, normally non-activating cytosolic calcium levels and ultimately pathologic cardiac muscle dysfunction^10–12^. Restoring calstabin2 binding using Rycals, a novel small-molecule class of drugs developed in our laboratory, has been shown to stabilize the channel and prevent SR calcium leak^13^.

Although historically four CPVT mutation hotspots have been identified (I exons 3-15, II 44-50, III 83-90, and IV 93-105)^14^, advances in sequencing technology have allowed for cost-effective complete screening of RyR2’s 105 coding exons^15^. As a result, HRS/EHRA expert guidelines recommend this comprehensive genetic test for any patient with suspected CPVT, along with cascade screening in family members following identification of the CPVT-causative mutation^16^. However, as genetic screening has become more readily available, it has been discovered that approximately 3% of ostensibly healthy individuals who are screened for RyR2 mutations carry a rare protein-altering variant of RyR2^7^, creating genetic noise that obfuscates interpretation of positive test results^14^. These ambiguous variants, or variants of unknown significance (VUS), introduce a “genetic purgatory” for the physician: a clinical dilemma in managing a family whose variant is not well studied^17^. This phenomenon is exacerbated when considering treatment recommendations such as implantable cardiac defibrillator (ICD) placement in the context of potentially fatal arrythmias^17^. Thus, there is an essential need for research focused on distinguishing pathogenic CPVT-causing variants from rare variants of inconsequential significance.

Although CPVT has become recognized as a significant cause of sudden cardiac death in children and young adults during the era of molecular biology^18, 19^, there is a large variability in treatment^20^. Furthermore, near-fatal event rates even in patients on beta-blockers remain unacceptably high^20^. With increasing evidence suggesting that beta blocker efficacy may be mutation-specific^21, 22^, it becomes imperative that treatment regimens that are more mutation-specific are developed.

The present study was designed to create an extensive database of RyR2 mutations associated with CPVT that will: 1) validate the current understanding of CPVT through analysis of the largest compilation of variant data based on comprehensive mining of the literature; 2) serve as a centralized, easily accessible data resource for physicians and patients to refer to for information on specific published RyR2 variants, including information about inheritance patterns, clinical manifestations, and treatment efficacies; and 3) form the foundation for future development of a machine learning-based pathogenicity predictor for VUS.

## Methods

### Study Selection and Data Extraction

Publication databases, including PubMed, Embase, and Scopus, were searched for articles written from their onset through October 2020. Keywords to narrow our search results were used, including, but not limited to, catechol* polymorphic ventricular tachycardia, channelopathic cardiac diseases, cardiac arrythmias, sudden cardiac death, syncope, and RyR2. A librarian at the Columbia University Medical Center Health Sciences Library developed, piloted, and executed the searches. English language studies reporting specific RyR2 variants associated with arrhythmias were included.

Data collected included but was not limited to, demographic information, age of CPVT symptom onset, cardiac history, treatment, and efficacy of treatment. Two investigators working independently assessed the quality of each article and extracted the data. Reasons for exclusion were recorded and cross-checked for agreement. Disagreements in exclusion reasoning or discrepancies in data extraction were resolved by discussion and mutual consensus. The database was designed primarily to include binary data to facilitate future analyses. Covidence software was used for the organization and tracking of the literature review.

### Structural Mapping

The human RyR2 cryo-EM model published in Miotto et al. was used as the base structure upon which we modeled our point mutations^11^. Point mutations were substituted into the wild-type structure, and final structure figures were created using ChimeraX, according to software instructions^23^.

### Statistical Analysis

When applicable, categorical data were compared using chi-squared or Fisher exact test and quantitative data were evaluated using one-way analysis of variance (ANOVA). *Post hoc* analyses were performed using Tukey’s HSD test. Statistical analyses were performed using R software (version 4.2.1; R Project for Statistical Computing, Vienna, Austria).

## Results

### Patient Demographics and RyR2 Database

Our search across the three platforms yielded 4510 articles, 1940 of which were excluded as duplicates. The remaining 2571 articles were screened by title and abstract, and 1688 were excluded (see **Supplemental Table 1**). 883 full texts were reviewed, and 227 were ultimately utilized in the final data extraction (**Supplemental Figure 1)**. Demographics are summarized in **Table 1**. Of the 1330 patients, 635 were female and 523 were male, 54.8% and 45.2%, respectively. Most patients inherited their RyR2 mutation (67.4%), and the overwhelming majority of the mutations were missense point mutations (93.8%).

**Table 1.** RyR2 Database Demographics.

|  |  |  |  |
| --- | --- | --- | --- |
| **Total patients** | 1330 | **Inherited RyR2 mutation\*\*\*\*** | 201 (67.4%) |
| **Average age of onset (years)\*** | 13.4 | **Missense mutation#** | 1248 (93.8%) |
| **Male\*\*** | 523 (45.2%) | **Deletion mutation#** | 59 (4.5%) |
| **Female\*\*** | 635 (54.8%) | **Insertion mutation#** | 5 (0.4%) |
| **Experienced sudden cardiac arrest\*\*\*** | 351 (35%) | **Nonsense mutation#** | 3 (0.2%) |

\* Reported for 513 patients (38.6%), \*\* reported for 1158 patients (87.1%), \*\*\* reported for 1002 patients (75.3%), \*\*\*\* reported for 298 patients (22.4%), # reported for 1315 patients (98.9%).

The compiled database contains 1330 individual cases of patients who carry heterozygous RyR2 mutations, of which 1008 (75.8%) had a primary diagnosis of CPVT or CPVT-like phenotype (**Figure 1A**). The patients’ age at diagnosis was skewed to the left, with the majority of cases being diagnosed before age 30 (**Figure 1B).** Data collected for each patient included specific RyR2 mutation, inheritance pattern and family history, cardiac symptomatology, treatment plans, and efficacy, among others. The entire database (CPVT and non-CPVT patients included) can be found attached as **Supplemental Table 2.**

**Figure 1.**
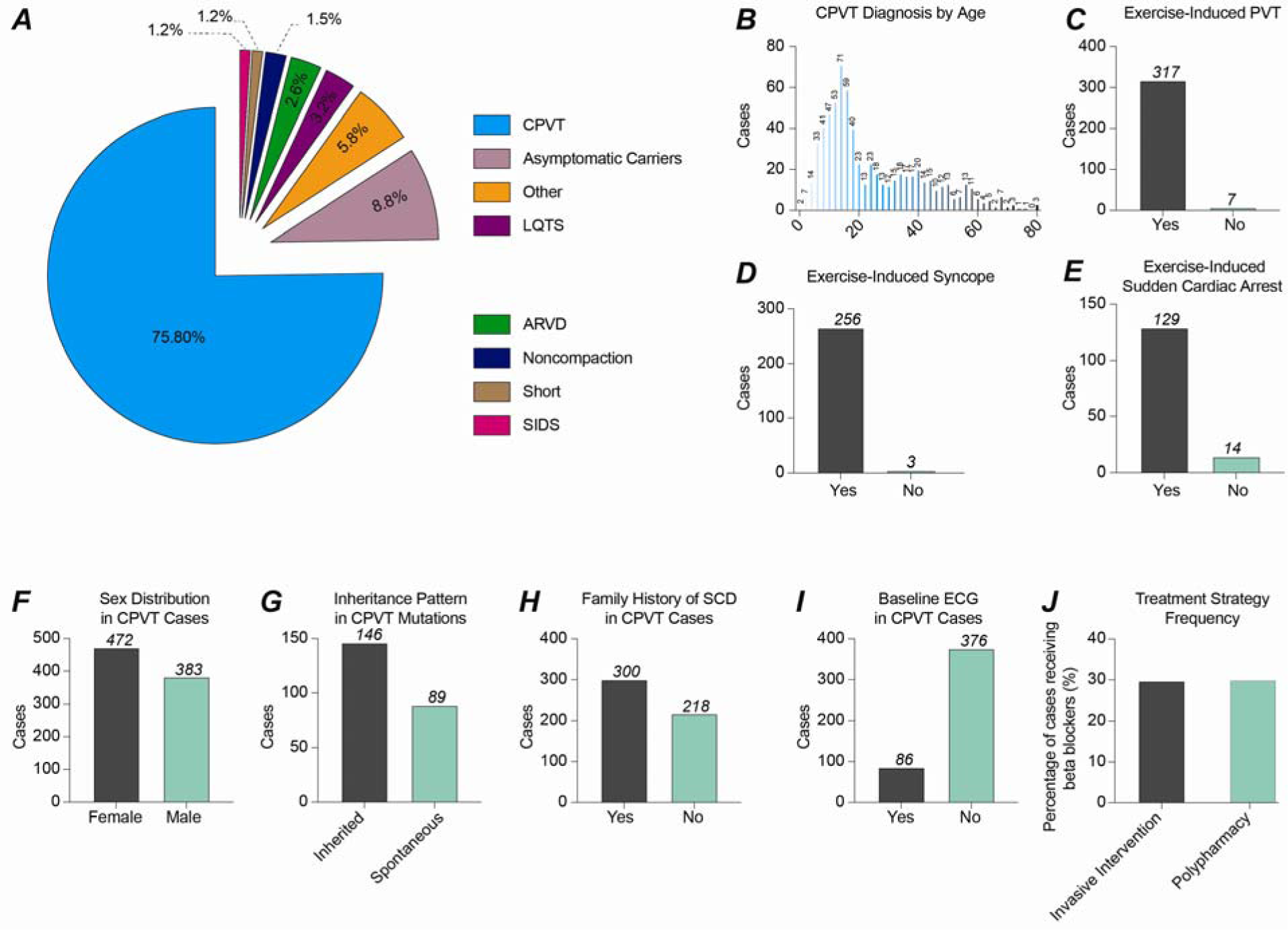
Clinical characteristics of RyR2-mutation-carrying CPVT patients. **(A)** Percentage of RyR2 mutations from the database that are associated with indicated diseases. 75.8% of mutations were associated with symptomatic CPVT, while 8.8% were found in asymptomatic carriers. **(B)** Number of CPVT cases diagnosed at each age. **(C)** Number of CPVT cases with exercise-induced polymorphic ventricular tachycardia (PVT). **(D)** Number of CPVT cases with exercise-induced syncope. **(E)** Number of CPVT cases with exercise-induced sudden cardiac arrest (SCA). **(F)** Distribution of CPVT cases in males and females. **(G)** Distribution of inherited vs. spontaneous CPVT RyR2 mutations. **(H)** Number of CPVT cases with a positive family history of sudden cardiac death (SCD). **(I)** Number of CPVT cases with baseline (resting) ECG changes. **(J)** Distribution of multi-faceted treatment approach in patients on beta-blockers.

### Clinical Characteristics of CPVT Patients

The analyses from here on are for cases with a primary diagnosis of CPVT or a CPVT-like phenotype (n = 1008). The vast majority of CPVT patients had structurally normal hearts (94.7%). Many patients underwent Holter ECG monitoring and/or cardiac stress tests as part of their diagnostic workup. Of those patients, 97.8% of documented polymorphic ventricular tachycardia (PVT) occurred in patients under exercise/stress conditions, highlighting the situation-dependent nature of CPVT **(Figure 1C)**. 98.8% of syncope was exercise/stress induced, as was 90.2% of sudden cardiac arrest (SCA) (**Figure 1D and 1E**, respectively).

Of those patients with reported sex (n=855), 472 (55.2%) were female and 383 (44.8%) were male (**Figure 1F).** The majority of CPVT patients had a confirmed inherited mutation (62.1%), with 57.9% of patients having a positive family history of sudden cardiac death (**Figure 1G and 1H, respectively).** 18.6% of patients had baseline ECG abnormalities, including bradycardia **(Figure 1I).**

### Therapeutics

The treatment for CPVT is multifaceted and often requires a poly-pharmaceutical approach to effectively manage the arrhythmia. In patients with detailed treatment records, 72.9% took beta-blockers; of these patients, 30.0% required additional rhythm-related medications such as flecainide, verapamil, or propafenone. Furthermore, 29.7% of the cases that received treatment with beta-blockers underwent an invasive therapeutic strategy such as ICD placement or left cardiac sympathetic denervation (**Figure 1J)**. When comparing therapeutic strategies between the most common mutations, a significantly greater proportion of those with the R420W (70.6%) required a poly-pharmaceutical approach compared to R420Q (8.3%, p = <0.001) in the setting of similar ICD usage. When comparing the two other most common mutations that occur in different regions of the protein, S2246L and G357S, cases with the S2246L mutation were significantly more likely to utilize a poly-pharmaceutical approach and to receive ICD placement (p < 0.001, p = 0.043, respectively) **(Supplemental Table 3)**. Lastly, of cases that reported the success of a medication, flecainide was the most effective and most frequently used in cases with an R420W mutation (91.67%, n = 12) compared to R420Q (0%, n = 2), S2246L (66.67%, n = 3) and G357S (n = 0).

### CPVT RyR2 Mutation Distribution

There are 286 unique RyR2 mutations recorded in the database, but mutations did not occur with equal frequency. G357S, R420Q, R420W, and S2246L were the most frequently reported RyR2 point mutations associated with CPVT (**Figure 2A)**. The deletion mutation N57_G91 was also frequently reported to cause CPVT. It is worth noting that in the case of the G357S mutation, the frequency was influenced by a large study that included genetic data from multiple generations of a single family, thus inflating its prevalence in our database beyond that of the general CPVT population. CPVT-associated mutations were also unequally distributed amongst exons, with exons 14, 46-47, and 90 each having at least 10 or more unique mutations **(Figure 2B).**

**Figure 2.**
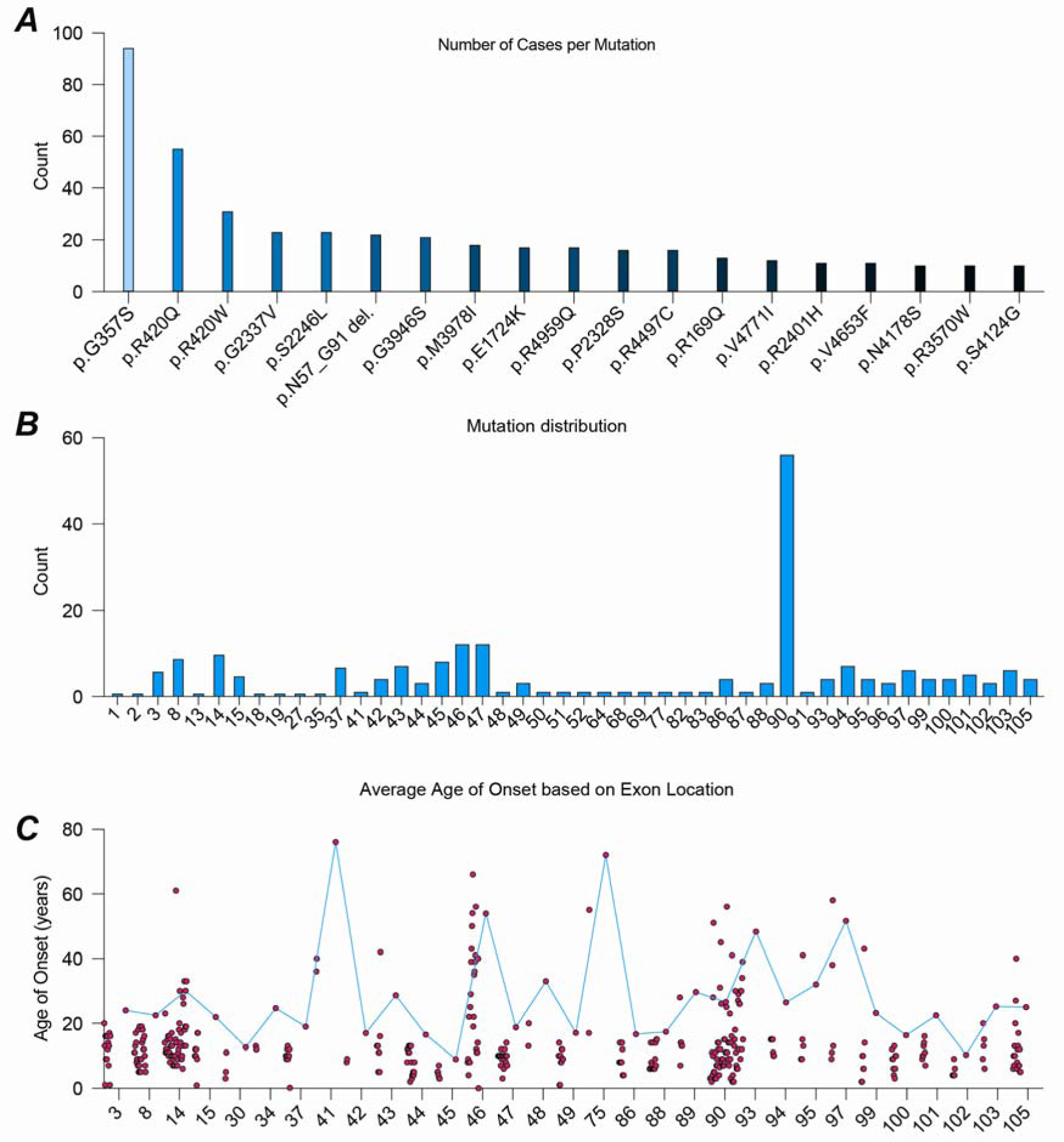
RyR2 mutation frequency in CPVT cases. **(A)** Number of cases with mutations frequently represented in the database. There are 286 unique RyR2 mutations recorded in the database, but mutations did not occur with equal frequency. G357S, R420Q, R420W, and S2246L were the most frequently reported RyR2 point mutations associated with CPVT **(B)** Number of unique mutations per specific exon. CPVT-associated mutations were also unequally distributed amongst exons, with exons 14, 46-47, and 90 each having at least 10 or more unique mutations. **(C)** Average age of CPVT symptom onset based on exon location.

### Structural mapping and effects on RyR2 domains and function

On a larger scale, patient mutations were also classified based on which exon and domain of the channel they affected. We used these data to determine whether a mutation in one region conferred a more severe phenotype over a mutation in another region, proxied by age of onset. The average age of onset for all CPVT patients, 13.4 years, showed great variability with a standard deviation of 10.6 years. When comparing all exons with more than one case, there was a significant difference in age of onset seen between groups (P < 0.001, **Figure 2C**). Of exons with more than one case, exon 45 had the lowest age of onset with a mean of 4.5 years (± 0.5, n = 6), and exon 41 had the highest with an average age of onset of 38 years (± 2.8, n = 2). Of the exons with a significant number of cases, exon 46 had the greatest variation (18.28 years, n = 26). Other exons—with more than one data point—that had an average age of onset below 10 were: 45, 102, 96, 30, 100, 44, 86, 42, 49, 88, 47, and 37 (**Figure 2C**).

We analyzed the historical CPVT mutation hotspots in the context of the atomic model: I (amino acids 44–466); II (2246–2534); III (3778–4201); and IV (4497–4959) (**Figure 3A and B**). We noticed that those hotspots are clustered in the core of the structure (colors), suggesting that the outer region of the cytoplasmic shell (grey) has less influence on CPVT.

**Figure 3.**
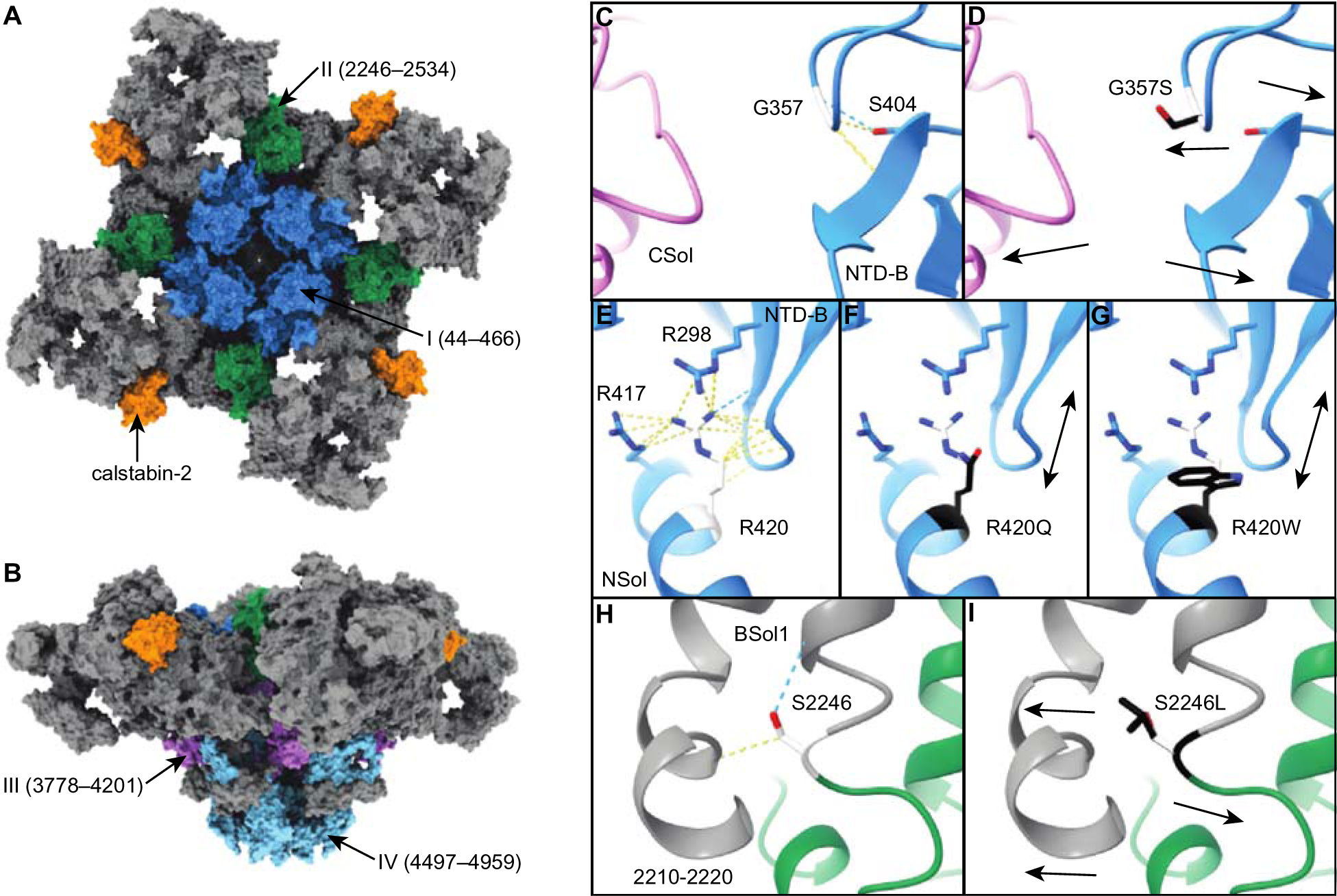
Structural changes caused by CPVT mutations in RyR2. **A-B**. Atomic model of RyR2 (PDB: 7UA5) from the top **(A)** and side **(B)** views, showing the CPVT hotspots: I (blue), II (green), III (purple), and IV (cyan). Residues outside the hotspots (grey) and subunit calstabin-2 (orange) are also shown. **C, E, H.** Atomic model of RyR2 focused on residues G357, R420, and S2246 (white). Calculated interaction contacts (yellow) and hydrogen bonds (blue) are shown as dash lines. **D, F, G, I.** Substitute residues S357, Q420, W420, and L2246 (black) were overlapped to interpret probable consequences. Hypothesized structural changes are shown as arrows.

It has been proposed that CPVT mutations affect the interaction and dynamics between RyR2 domains. To better understand the effects of mutations on the structure and microdomain environments of the channel, we mapped the G357S, R420Q/R420W, and S2246L point mutations—which occur in high frequency—into the previously published high-resolution structure of wild-type RyR2 (**Figure 3C-I**). The mutation G357S probably disturbs the local β-turn secondary structure, stabilized by Gly residues, affecting the interaction within the NTD-B domain and the interaction between the NTD-B and the CSol domains (**Figure 3C and D**). The mutations R420Q and R420W probably disrupt the interaction network centered on R420, affecting the interaction between the NSol and NTD-B domains (**Figure 3E-G**). The different size, polarity, and hydrophobicity of the substitute residues (Q420 and W420) might have dissimilar effects on the flexibility of the NTD-B domain, causing different degrees of symptoms. Finally, the mutation S2246L, which also has a low age of onset, substitutes a small polar residue (Ser) by a large hydrophobic residue (Leu). This drastic change introduces steric impediments that most likely affect the intra-domain stability, increasing the domain flexibility **(Figure 3H and I)**. Interestingly, the three sites analyzed seem to operate by different local structural mechanisms but have similar global consequences presenting as loss of inter-domain interactions and increased flexibility. Indeed, we and others have previously reported that single channel and Ca^2+^ imaging studies of R420Q/R420W, and S2246L mutant RyR2 channels have shown that these mutations result in leaky channels^12, 24–26^, and knock-in mice with the S2246L, R420Q and R420W mutation have a CPVT phenotype^27, 28^.

The RyR2 structure can be subdivided into protein domains and subdomains (**Figure 4A and Supplemental Table 4).** Differences in age of onset based on protein domain and based on protein subdomain were also significant (p = 0.032, p < 0.001) (**Figures 4B and 4C**). The junctional solenoid domain (JSol) had the lowest age of onset with the lowest variation (9.5 years ± 3.6), which was significantly different on post hoc analysis from the age of onset for cases with mutations in the Shell-core linker peptide (SCLP, 36.0 years ± 26.9, p = 0.038). The core solenoid domain (CSol), which showed a similar age of onset (10.9 years ± 8.6), was also significantly lower than the SCLP (p = 0.029). The finding that mutations in these domains are the most aggressive is not surprising because the JSol and CSol domains are signal transduction domains, connecting the regulatory cytoplasmic domains with the transmembrane domain. Mutations in these domains would likely have more impact on channel regulation than mutations in the cytoplasmic domains. When evaluating the subdomains of RyR2, JSol still had the lowest age of onset. Subdomain post hoc analysis showed a significantly lower onset age in the pore region (9.9 years + 7.6) compared to SCLP (36.0 years + 26.9, p = 0.031), and the difference in onset age approached statistical significance when comparing NTD-A to SCLP (p = 0.064).the finding that mutations in this subdomain are severe is not surprising because mutations in the pore subdomain would directly affect pore opening dynamics resulting in leakier RyR2. Finally, the SCLP, which contains the calmodulin binding peptide, showed the highest age of onset in both analyses. This suggests that dysregulation by calmodulin is less likely to result in CPVT than mutations that affect the structure and dynamics of RyR2.

**Figure 4.**
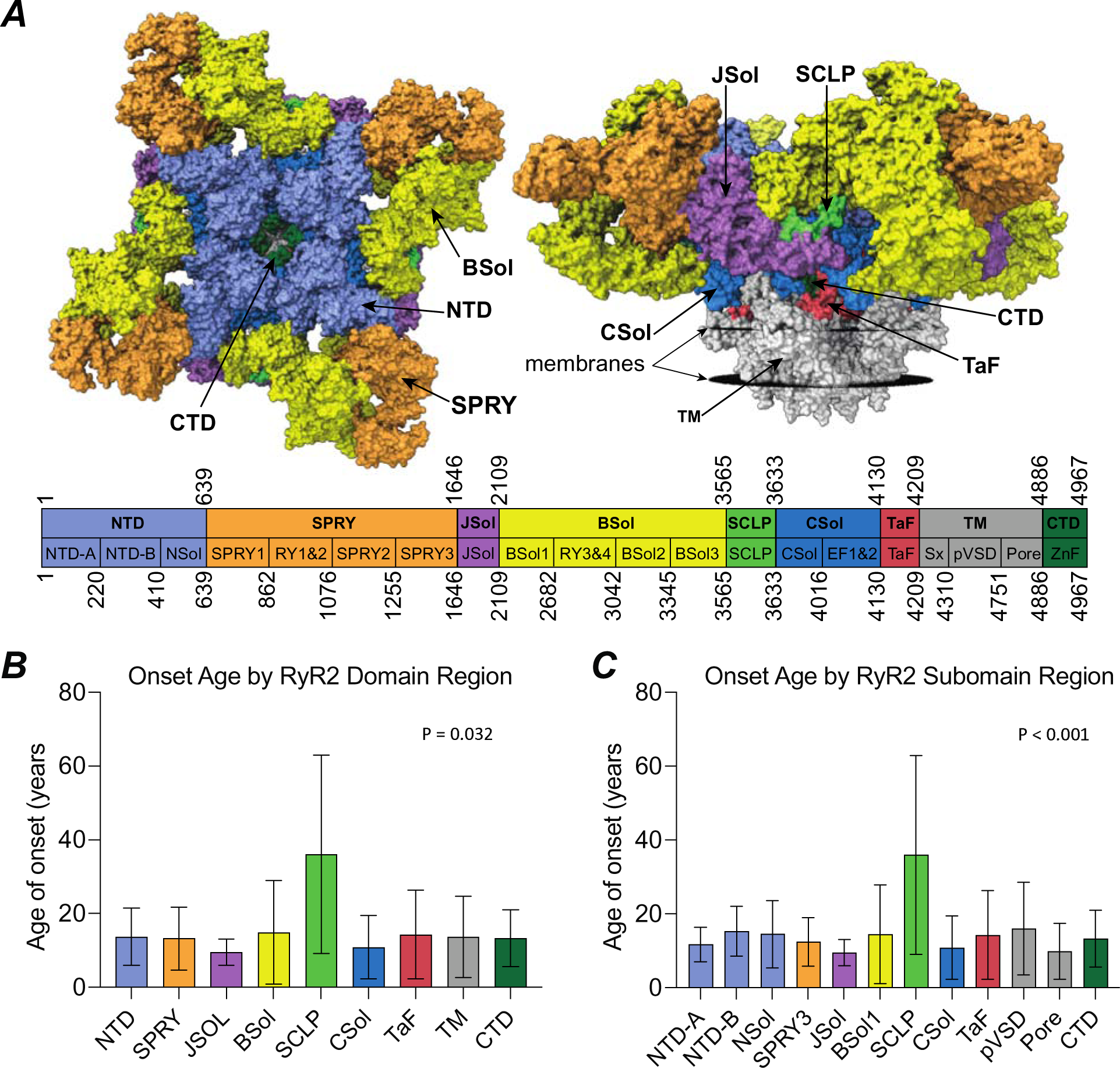
Age of onset of CPVT symptoms based on RyR2 Domain and Subdomain location of mutation. **(A)** RyR2 delineated domains and subdomains **(B)** Average age of onset by RyR2 domain. **(C)** Average age of onset by RyR2 subdomain. Subdomains with only one case were excluded.

## Discussion

In this study, we generated a database of CPVT-associated RyR2 mutations to facilitate the characterization of mutation phenotypes and elucidate trends in the data that could help guide future diagnosis and treatment of CPVT patients. We compiled the largest and most comprehensive database of RyR2 CPVT mutations, to our knowledge, including as much genetic and clinical information for each case as was available. Using these data, we were able to characterize the prevalence of certain hallmark symptoms of CPVT. In assessing the inheritance patterns of the patients’ mutations, most were inherited and had a positive family history of sudden cardiac death; however, we found that a greater proportion of patients with spontaneous mutations experienced sudden cardiac arrest compared to those with inherited. For this reason, it is important to consider genetic testing in young patients with suspicion for CPVT despite a lack of family history, as identifying and addressing a pathogenic mutation could prevent future cardiac events. It is important to acknowledge, however, that the difference in the incidence of sudden cardiac arrest between spontaneous mutation cases and inherited mutation cases is partially attributable to many cases who inherited a mutation being family members of an “index” case and thus, are medically managed before an event, compared to spontaneous cases who often are the first in the family to present with cardiac arrest.

The mutation distribution in our database aligns well with the previously defined four mutation hotspots discussed earlier, as well as Kapplinger et al.’s refined hotspot regions (exons 3, 8, 14, 43, 47–49, 81, 83, 88–90, 93, 95, 97–101, 103, 105)^16^. In addition to these exons, our database showed many unique mutations in exons 37 and 42; thus, we posit that these exons should be considered a part of hotspot II.

We collected data on mutation location and analyzed the phenotypic variability based on the domain, subdomain, and exon distribution. Age of onset of symptoms, typically syncope or sudden cardiac arrest, differed significantly at all three structural levels. Such patterns may have implications for genetic screening results and potential therapies. For example, relatives of an index case found to have a mutation in exon 45, which had a low age of onset, may require earlier intervention, more invasive management, or increased monitoring. Moreover, analyzing the clinical impact of mutations based on RyR2’s regions can facilitate the focused discovery of novel therapies.

Rycals are a novel class of small-molecule drugs that restore the binding of calstabin2 to RyR2 and prevent RyR2 mediated diastolic SR Ca^2+^ leak, thus normalizing Ca^2+^ homeostasis^13^. Rycals are an effective treatment for RyR2-associated diseases in pre-clinical models^29–31^.

In our database, the single case with a known age of onset who had a mutation in the calstabin2 binding region experienced his first symptoms at 8 years old. This early age of onset for a case with a mutation in a region in which preliminary studies have demonstrated effective targeted therapy use further demonstrates the importance of mutation regional analysis in identifying therapeutic targets and highlighting the potential use of age of onset as a proxy for relative pathogenicity.

The use of age of onset as a clinical proxy for relative pathogenicity is further corroborated by the three mutations with the earliest age onset in our database (G3946S, R169Q, and S2246L) being represented in Kapplinger et al.,’s list of “Near-Definite Pathogenic Variants”^14^. Mutations that have been functionally characterized showing evidence for leaky channels including RyR2-R2474S^32, 33^, RyR2-N2386I^33^, and RyR2-L433P^33^ also are located on exons frequently associated with an early age of onset. Finally, given that onset symptoms are usually syncope or sudden cardiac arrest, the clinical utility of this measure is significant.

The aggregation of a large quantity of data for many different RyR2 mutations makes the heterogeneity in phenotype severity between mutations apparent. For example, between two unrelated families with the M3978I mutation, every case was symptomatic and met clinical criteria for CPVT. In contrast, every G155R case in our database was entirely asymptomatic. As is expected with mutations reported more frequently in the literature, they have the greatest heterogeneity within the same mutation. For example, for R420Q, some cases experienced cardiac arrest, some were asymptomatic, and a few exhibited bradycardias. This increased variability is likely partially due to increased sample size compared to that of a rarer mutation such as G155R. It is also attributable to the survivability of that mutation; rarer mutations are likely to be more fatal and thus do not get passed down across generations. More detailed family trees are required to calculate the penetrance of specific variants; however, a tool for predicting pathogenicity for variants of unknown significance would provide even greater utility. Such a tool could be a machine learning model trained by comparing pathogenic and benign variants utilizing variables that include the biophysical properties of amino acid changes, the constraint of the location in the gene (utilizing a score such as pLI), and clinical data of patients with such variants. With the creation of such a tool in mind, this database was designed to facilitate the modeling process, especially as more data continue to be accumulated.

Almost a third of patients in our review did not respond adequately to beta-blockers, requiring additional anti-arrhythmic medications and/or an invasive therapies such as ICD placement or left cardiac sympathetic denervation. A significant portion of these patients were treated with flecainide. Liu et al. found that while flecainide did indeed prevent arrhythmias in a CPVT RyR2 mutant mouse model, it had no effect on intracellular calcium homeostasis^34^. These findings suggest that the antiarrhythmic role of flecainide in CPVT is likely primarily related to its classic mechanism of inhibiting sodium channels, preventing propagation of arrhythmias that may be triggered by aberrant calcium handling rather than inhibition of RyR2 itself. These findings underscore the need for personalized approaches depending on the mutation and improvement in treatment options in general—there remains a gap in addressing the underlying mechanism of the disease itself.

This calls for the introduction of new therapies more specific to RyR2 and its interactions with surrounding proteins. In 2019, Park et al. highlighted calmodulin-dependent protein kinase II activation as a key event in arrhythmia formation in patients with CPVT, identifying a target that could be inhibited to potentially treat RyR2 associated CPVT^35^. Similarly, as mentioned previously, Rycal’s mechanism of stabilizing RyR2 in the closed state suggests that these drugs will be a promising option to address the channel dysfunction in all CPVT patients and thus could result in measurable clinical benefit ^13^.

The size and depth of this database make it a useful tool to educate patients, inform physicians, and potentiate research regarding CPVT and its treatment. Compared to other available data of CPVT patient cases, our database encompasses a broader scope of medical, genetic, and structural information, allowing an analysis of both the clinical and scientific aspects of each CPVT mutation. Furthermore, our analyses of the structural effects of certain mutations illustrates the structure-function relationship of RyR2 and begins to narrow the gap in the understanding of the underlying pathogenicity of less robustly studied CPVT mutations. For physicians, this database can facilitate gathering data on rare variants directly from literature to help guide management based on patient presentation, genetic findings and treatment options. Using the data in this database, as well as the easily accessible literature related to each mutation, healthcare professionals and patients can make more informed decisions based on specific RyR2 mutations with the hope of decreasing the time to diagnosis and increasing treatment efficacy.

Taken together, this project has produced, to our knowledge, the most comprehensive compilation to date of CPVT-associated RyR2 mutations and their associated clinical phenotypes. Data were analyzed to determine the frequency of the most salient clinical features of CPVT and the efficacy of currently available treatment options. Finally, this study has important clinical relevance because it provides a potential roadmap to risk stratify CPVT patients based on the location of their RyR2 mutation.

## Data Availability

All data produced in the present work are contained in the manuscript

## Acknowledgments

Funding: The work was supported by grant NIH R01HL145473, R25HL156002, P01 HL164319 T32 HL120826.

## Competing interests

Columbia University and ARM own stock in ARMGO, Inc., a company developing compounds targeting RyR and have patents on Rycals.

## Author contributions

LS, HB, ARM designed the study, LS, HB, AC, GW, JA, MM, CM, YL, ZD, HD, SR performed literature review, analyzed data, and LS, HB, SR, HD, ARM wrote/edited the manuscript.

## Supplementary Appendix

This appendix has been provided by the authors to give readers additional information about their work.

**Supplemental Figure 1.**
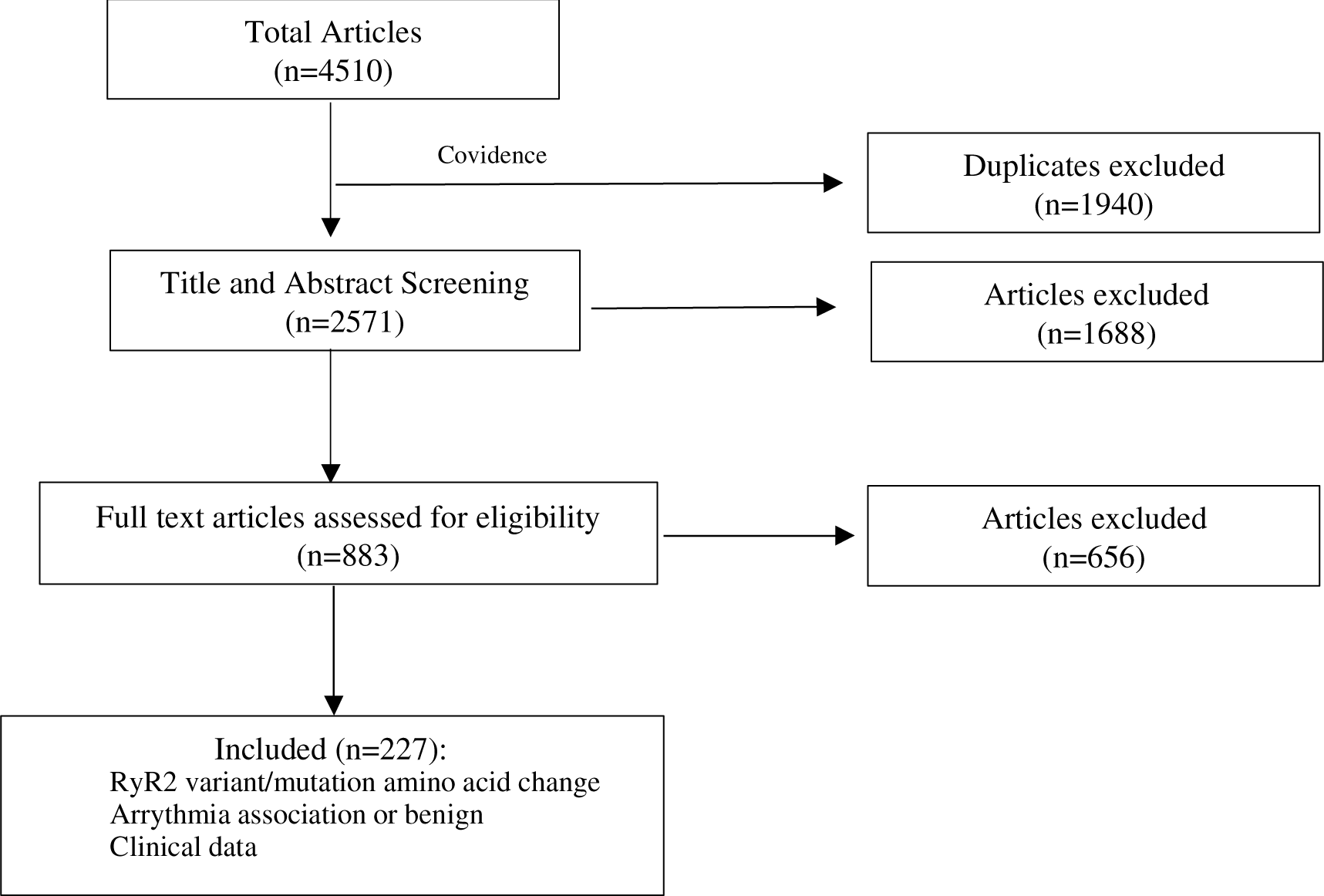
Literature review workflow. Search yielded 4510 articles; 1940 duplicates removed. 2571 titles/abstracts screened; 1688 excluded. 883 full texts were assessed: 656 were excluded. 227 articles were included in final data extraction.

**Supplemental Table 1.** Inclusion and Exclusion Criteria.

|  | **Inclusion Criteria** | **Exclusion Criteria** |
| --- | --- | --- |
| **Title/Abstract**<br>**Screening** | - Clear indication that a variant/mutation of RyR2 is investigated in the paper<br>- Abstract suggests there is a possibility of clinical data presented from the patient harboring the RyR2 variant discussed in the paper | - Main focus of the paper is alterations in pathways associated with RyR2, but not changes to RyR2 itself<br>- Heart failure unrelated to CPVT |
| **Full Text**<br>**Screening** | - Variant/mutation of RyR2 explicitly stated<br>- Clinical data (must have one of the characteristics below):<br>- Arrhythmia diagnosis associated with RyR2 variant/mutation<br>- If no diagnosis, but clinical suspicion, must have basic CPVT clinical symptom(s) at presentation<br>- Case with RyR2 variant/mutation and no cardiac symptoms | - Clinical cases associated with heart failure unrelated to CPVT<br>- CPVT or CPVT-suspected cases with variant/mutation of gene besides RyR2 in addition to having an RyR2 mutation<br>- E.g., if RyR2 and CASQ2 are both mutated in the same patient<br>- Cannot distinguish or identify individual clinical data |

**Supplemental Table 3.** Mutation specific treatment strategy. Demonstrates frequency of utilization of other therapeutic strategies in addition to beta-blockers for the most common mutations.

| MUTATION | POLY-PHARMACEUTICAL<br>APPROACH | ICD | TOTAL NO. RECEIVING<br>BETA-BLOCKERS |
| --- | --- | --- | --- |
| R420W | 12 (70.6%) | 7 (41.2%) | 17 |
| R420Q | 2 (8.3%) | 13 (54.2%) | 24 |
| S2246L | 4 (40%) | 8 (80%) | 10 |
| G357S | 1 (1.1%) | 41 (43.6%) | 94 |

**Supplemental Table 4.**
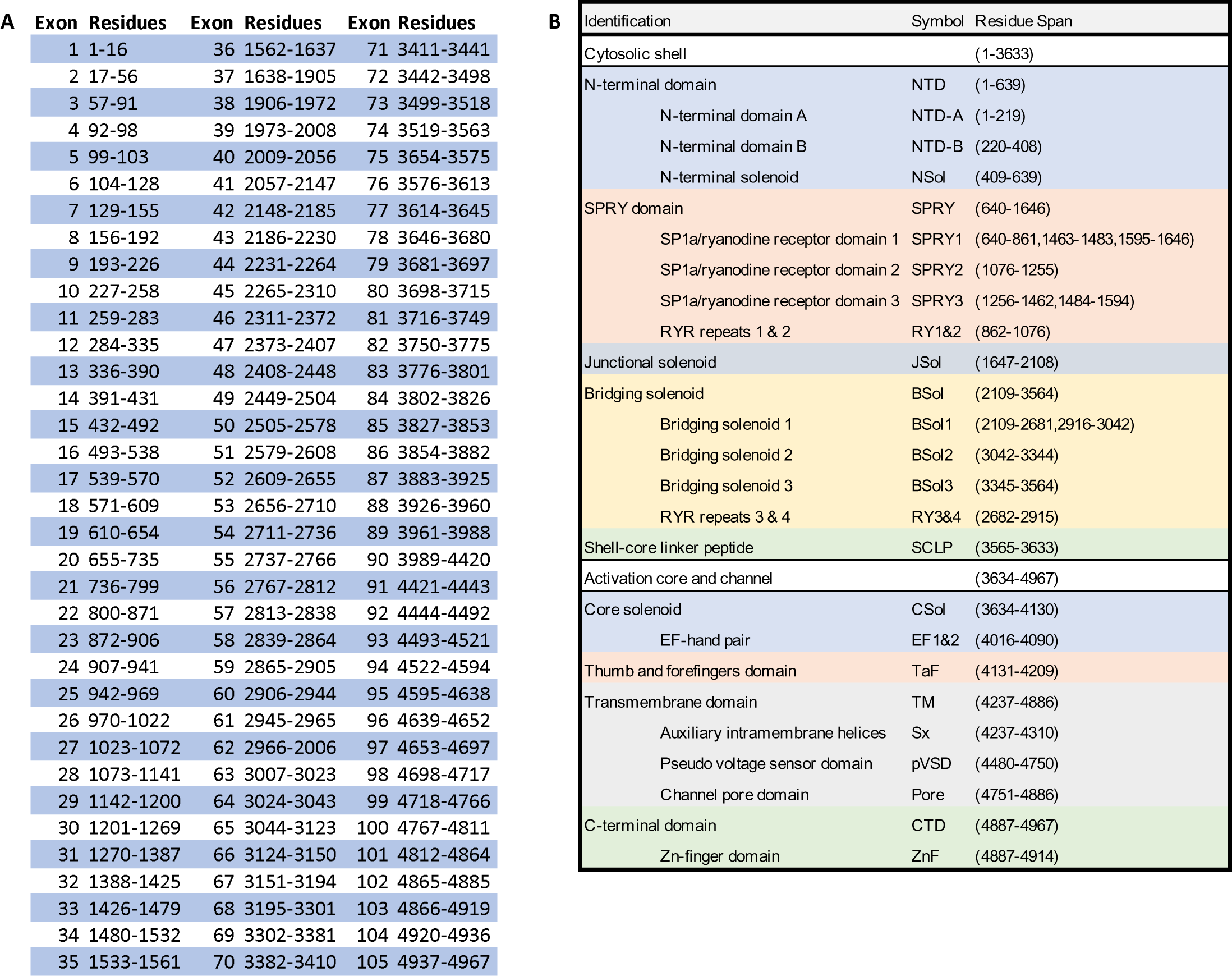
Exon, Domain and Subdomain residue alignment.

## Notes

### Author Declarations

Source of data are publicly available on pubmed.

